# Impact of control selection strategies on GWAS results: a study of prostate cancer in the UK Biobank

**DOI:** 10.1101/2025.10.08.25337574

**Authors:** Jingzhan Lu, Johan H. Thygesen, Robin N. Beaumont, Michael N. Weedon, Harry D. Green

## Abstract

As GWAS studies move from array-based genotyping to whole exome and genome sequencing, there is a significant increase in cost. Applying an appropriate technique for the selection of which controls to include, in large studies where more potential controls are available than needed for the study, may be a useful technique for minimising resource intensity while maintaining statistical power. We evaluated three control selection strategies in prostate cancer GWAS using 15,250 UK Biobank cases: (a) all controls, (b) matched controls, and (c) random selection. Both (b) and (c) achieved comparable power in detecting significant loci relative to (a), but matched controls (b) showed greater consistency in identifying leading SNPs. However, using (b) matched controls reduced discovery power by ∼30% compared with (a) all controls, highlighting a trade-off. Matching controls (1:4 ratio) offers a cost-effective approach for targeted SNP analysis across phenotypes but may miss novel associations.

**Availability and Implementation:** R code for implementing matching and random control selection is provided and available on GitHub (https://github.com/Jingzhan-Lu/GWAS-Control-Selection).

## 1. Introduction

### 1.1 Genome-wide association studies

Standard disease GWAS studies are typically case-control studies aimed to detect genetic variation, known as single nucleotide polymorphisms (SNPs), which associate with the disease of interest. Significant associations between SNP and disease can help understand aetiology and potentially contribute to risk stratification tools (Wu et al., 2010). Also, within cancer research, GWAS has been used successfully to identify over 200 genetic risk loci associated with susceptibility to prostate cancer to date (Conti et al., 2021; Qian et al., 2023).

### 1.2 Control selection further complicated in large longitudinal study

Chatterjee et al., (2016) emphasized that the validity of case-control studies relies heavily on the appropriate selection of controls. Proper control selection reduces the risk of selection bias and enhances the credibility of study findings. This issue is particularly salient in genome-wide association studies (GWAS) using biobank-scale datasets, where vast numbers of potential controls are available, making the strategy for selecting appropriate controls a critical study design consideration.

As digital health infrastructure expands and increasing volumes of electronic health records (EHRs) and biobank-based data become accessible, the challenge of selecting appropriate controls becomes more complex. Prostate cancer is a highly heritable disease, with heritability estimates as high as 58% (95% CI, 51%–63%) according to twin studies (Mucci et al., 2016). In this study, we will use prostate cancer as an example phenotype because there is a large genetic component to prostate cancer. In the UK Biobank, once prostate cancer cases are defined, virtually any other individual may qualify as a “healthy” control. Therefore, instead of including all eligible controls, we limited our selection to a maximum 1:4 ratio (Hong & Park, 2012), aiming to balance power, reduce bias, and optimise computational efficiency. This study investigates how different strategies for control selection impact GWAS outcomes, with an emphasis on sufficient statistical power and cost-benefit of time and computational resources.

In large-scale cohorts, it is common practice to include all available non-case individuals as controls in GWAS. This issue is especially critical in population-based studies where subtle ancestral differences may remain even after statistical adjustments, leading to both false-positive and false-negative associations (Armstrong et al., 2014; Uffelmann et al., 2021). Failure to adequately address this issue remains a key limitation in using large biobank or EHR data where extensive inclusion of all controls without careful matching may compromise the accuracy of GWAS results.

### 1.3 Cost and scalability challenges in case–control GWAS

Traditionally, GWAS studies have been performed on SNP array data imputed to ∼10 million variants, whole genome sequencing (WGS) and whole exome sequencing data (WES) are now more readily available in large cohort datasets such as the UK Biobank (Bycroft et al., 2018) and All of Us (“The ‘All of Us’ Research Program,” 2019). While WGS data increases the potential to find novel rare variants, the increased scale of the data causes an increase in computational intensity. Most research in this field is now performed on a cloud-based research analysis platform (RAP) in which researchers pay for the computing resources used, so computational costs also incur a financial cost.

More critically, when conducting new case–control studies, it is not sufficient to sequence only the cases; appropriately matched controls must also be identified and sequenced, which often represents one of the most expensive components of the study. For example, even within large biobank resources, identifying around 3,000 prostate cancer cases for whole-genome sequencing also requires sequencing an equivalent or larger number of unaffected male controls. Selecting an appropriate number of individuals and defining a suitable health control cohort can decrease sequencing costs and logistical complexity. Against this backdrop, case-control selection could potentially reduce cost while minimising power loss, especially for studies planning to run multiple GWAS across many phenotypes.

### 1.4 Matching Strategy for Control Selection

Matching is a commonly employed strategy in case-control studies to ensure comparability between cases and controls by balancing important covariates, thereby minimising the influence of confounders on case-control study results. Several matching techniques exist, including nearest-neighbour matching, optimal matching, and ratio-based matching. In a comprehensive evaluation using Monte Carlo simulations, Austin (2014) compared 12 propensity score matching algorithms, such as optimal matching, greedy nearest-neighbour matching without replacement, and calliper matching, which reported that nearest-neighbour matching consistently demonstrated superior performance. Following these findings, this study, applied a propensity score–based nearest-neighbour matching to select controls with characteristics similar to cases.

## 2 Methods

### 2.1 UKBB genotyping quality control (QC) and DNAnexus

The UK Biobank (UKBB) genotyping data includes 488,377 participants. All analyses were performed within the secure UKBB Research Analysis Platform (hosted on DNAnexus). The quality control (QC) procedures were based on the publicly released metrics and recommendations provided by the UKBB research team. Samples with a mismatch between self-reported sex and genetically inferred sex were excluded, as were individuals of non-European (non-CEU) ancestry. In addition, 968 outliers based on heterozygosity or missing genotype rate were removed, and samples with sex chromosome aneuploidy were excluded. Variant-level quality control excluded genetic variants with an imputation INFO score <0.7. Minor allele frequencies (MAF) were recalculated within the prostate cancer phenotype subset, and variants with MAF < 1% were excluded.

### 2.2 UKBB phenotypes and covariates definition

Prostate Cancer cases (N = 15,250) were identified in the UK Biobank using cancer registry records based on the ICD-10 code C61, any participant with this diagnostic code was considered a case. Female participants inferred by genetics were excluded from the analysis. A total of 208,128 male participants without a diagnosis of prostate cancer were available as potential controls. A total of 13 covariates (recruitment age, assessment centre, genotyping chip and first 10 principal components) were used in the GWAS analysis and matching process.

### 2.3 GWAS power calculations

A sample size with sufficient statistical power is critical to the success of genetic association studies to detect causal genes of human complex diseases. We performed a power calculation for a case-control GWAS with 15,250 prostate cancer cases and 4 times the number of controls (1:4 case-control ratio). Power calculations were conducted using the developed Genetic Power Calculator framework on additive genetic models (Green, 2025), the key parameters included varying minor allele frequencies (MAF) from rare to common variants, and odds ratios (OR) of 1.5, 2, and 2.5, reflecting small to moderate genetic effect sizes. This power calculation reflects the theoretical baseline under random control selection, against which the observed gains from random controls can be evaluated.

### 2.4 Three Control Selection strategies and GWAS analysis

In this study, we conducted GWAS for prostate cancer within the UKBB using three different control selection strategies to evaluate how the different control definitions affect GWAS results. All controls, included all male participants from the whole population without prostate cancer (N = 208,128); Match controls, selected controls matched to cases based on 13 covariates; and Random controls, included a random selection of male controls. For both strategy match and random, a 1:4 case-to-control ratio was used (n = 61,000). For each strategy, prostate cancer GWAS was performed independently using REGENIE software. To assess the impact of control selection on GWAS results, we compared our findings against the largest-to-date trans-ancestry meta-analysis of prostate cancer (Conti et al., 2021), which included 107,247 cases and 127,006 controls. We used the 269 genome-wide significant SNPs reported in Conti et al. (2021) as a benchmark, matching them to corresponding SNPs in our GWAS results to evaluate the consistency and effect size of our GWAS results.

### 2.5 Nearest Neighbour Matching and Random for Control Selection

To minimize confounding and ensure balance in key covariates between prostate cancer cases and controls, we applied nearest-neighbour propensity score matching using the MatchIt R package (v4.5.2). The nearest-neighbour algorithm was employed without replacement to pair each case with four control individuals having the most similar estimated propensity scores. Matched between cases and controls was assessed using empirical quantile-quantile (eQQ) plots to evaluate covariate balance. Post-matching, matched individuals were retained for downstream GWAS. For Random, we randomly selected four controls for each case from the eligible control population. To ensure reproducibility, a random subset of 61,000 male participants was selected from the full pool of 208,128 eligible controls using a fixed random seed (set.seed = 42).

### 2.6 Locus Signal Analysis

To identify independent association signals from GWAS summary statistics, we implemented a custom locus-filtering script written in Perl and LocusZoom (Pruim et al., 2010). The script processes one or more GWAS result files and extracts lead variants based on statistical significance, variant quality, and genomic proximity. For variants passing the filtered criteria, the script then performs locus-level clustering using a distance-based approach. Specifically, variants within ±500 kb of each other on the same chromosome were grouped as a single locus, and the variant with the most significant P-value (i.e., highest – log10P) within each group was selected as the lead SNP. This procedure enables efficient identification of genome-wide significant loci while accounting for local linkage disequilibrium and variant quality.

## 3 Results

We characterised the demographic and covariates of the GWAS cohorts to evaluate their comparability (**Table 1**). The full set of cases (N = 15,250) was contrasted with the three control strategies: all controls (N = 223,378), matched controls (N = 76,250), and random controls (N = 76,250).

**Table 1.** Baseline characteristics of UK Biobank participants included in prostate cancer case and control cohorts under different GWAS control selection strategies.

| Baseline characteristic | All cases<br>N=15,250 |  | All-controls*<br>(cases + controls)<br>N=223,378 |  | Match*<br>(cases + controls)<br>N=76,250 |  | Random*<br>(cases + controls)<br>N=76,250 |  |
| --- | --- | --- | --- | --- | --- | --- | --- | --- |
|  | n | % | n | % | n | % | n | % |
| Age |  |  |  |  |  |  |  |  |
| ≤40 | 14 | 0.09% | 2,555 | 1.14% | 69 | 0.09% | 773 | 1.01% |
| 40-49 | 562 | 3.69% | 49,255 | 22.05% | 2,934 | 3.85% | 14,855 | 19.48% |
| 50-59 | 3,514 | 23.04% | 71,261 | 31.90% | 17,278 | 22.66% | 23,241 | 30.48% |
| 60-69 | 11,001 | 72.14% | 99,108 | 44.37% | 54,972 | 72.09% | 36,924 | 48.42% |
| ≥70 | 159 | 1.04% | 1,199 | 0.54% | 997 | 1.31% | 457 | 0.60% |
| Sequencing Chip |  |  |  |  |  |  |  |  |
| 1- BiLEVE array | 1,710 | 11.21% | 24,946 | 11.17% | 8,594 | 11.27% | 8,560 | 11.23% |
| 2- Axiom array | 13,540 | 88.79% | 198,432 | 88.83% | 67,656 | 88.73% | 67,690 | 88.77% |
| Recruitment Centre |  |  |  |  |  |  |  |  |
| 11007-Reading | 1,050 | 6.89% | 13,203 | 5.91% | 4,657 | 6.11% | 4,531 | 5.94% |
| 11008-Manchester | 857 | 5.62% | 13,223 | 5.92% | 4,467 | 5.86% | 4,496 | 5.90% |
| 11009-Newcastle | 1,000 | 6.56% | 16,517 | 7.39% | 5,627 | 7.38% | 5,574 | 7.31% |
| 11010-Leeds | 1,443 | 9.46% | 19,723 | 8.83% | 6,762 | 8.87% | 6,746 | 8.85% |
| 11011-Bristol | 1,312 | 8.60% | 18,726 | 8.38% | 6,250 | 8.20% | 6,436 | 8.44% |
| 11013-Nottinham | 1,021 | 6.70% | 15,188 | 6.80% | 5,387 | 7.06% | 5,118 | 6.71% |
| 11014-Sheffield | 922 | 6.05% | 13,708 | 6.14% | 4,895 | 6.42% | 4,751 | 6.23% |
| 11016-Liverpool | 909 | 5.96% | 14,794 | 6.62% | 5,155 | 6.76% | 5,070 | 6.65% |

\* Total Sample size of cases + controls

We compared the demographic and covariates characteristics of all cases with the three GWAS cohorts (All, Match, Random). Among them, the match control selection showed the closest similarity to the case group, especially in age distribution (e.g., 72.1% aged 60–69 in both groups), sequencing centre proportion, and genetic principal components. These results indicate that match control selection method best preserved covariate balance relative to the case group.

### 3.1 GWAS power calculation and cost analyiss

To ensure sufficient statistical power for detecting genetic associations, we performed power calculations in **Figure 1**. for a prostate cancer GWAS using 15,250 cases and a 1:4 case-control ratio. Under an additive model, the study reaches over 80% power to detect variants with an odds ratio (OR) of 1.5 when the minor allele frequency (MAF) exceeds 0.007. For variants with ORs of 2 or higher, the study achieves nearly 100% power even at lower MAFs (as low as 0.003). These results demonstrate that a 1:4 case-control ratio using random controls provides sufficient GWAS power and can be generalised to other phenotypes requiring large-scale genetic association studies.

**Figure 1.**
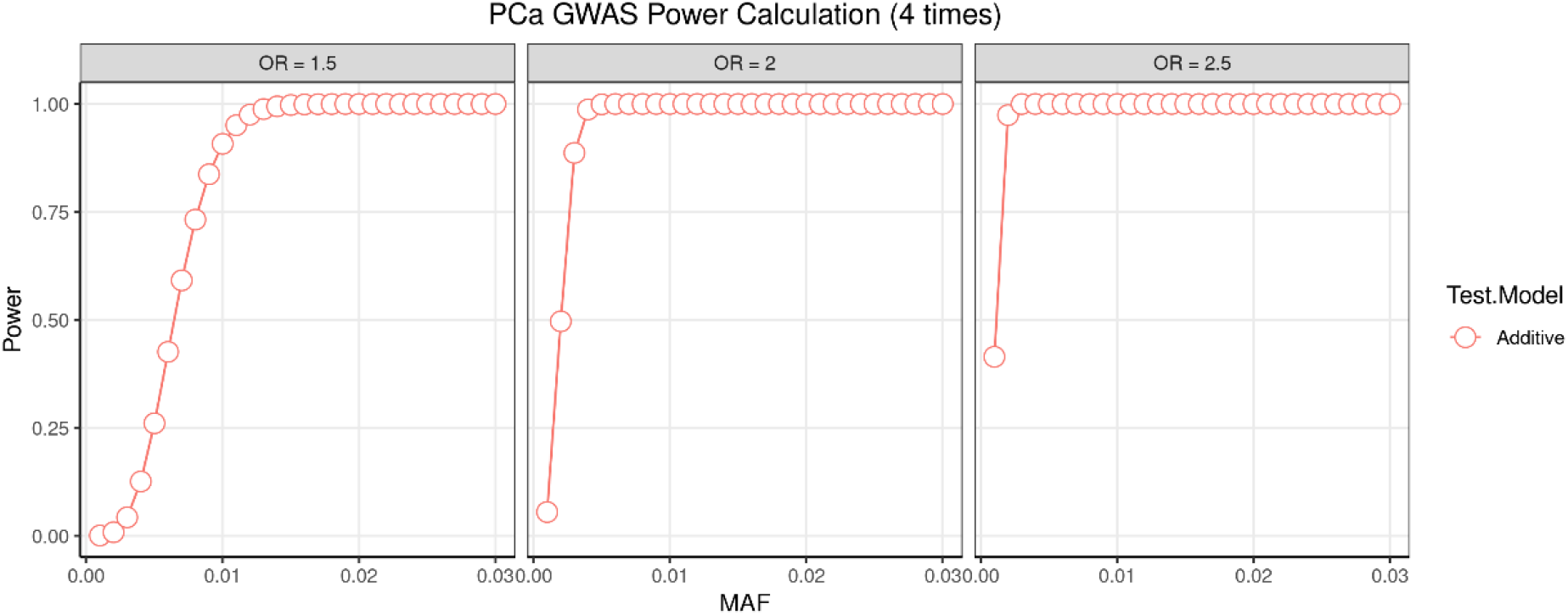
Power estimation for prostate cancer under a 1:4 case-control design. Three panels show additive model power curves for odds ratios (OR) of 1.5, 2, and 2.5. The x-axis represents minor allele frequency (MAF) and the y-axis shows statistical power.

Using a case-control selection strategy reduced the total sample size from 200,000 to 60,000, theoretically offering a 70% reduction in computing resource usage, or 91% reduction for algorithms are of O(N^2) complexity. Sample size reduction and thus resources saved will depend heavily on the prevalence of the phenotype and case/control ratio used. This offers both a cost-saving opportunity and an environmental benefit from reduction in high performance computing usage.

### 3.2 Manhattan Plots of GWAS results

In Figure 3, the Manhattan plot revealed genome-wide significant loci peaks are consistent with previously reported prostate cancer susceptibility regions. These consistent patterns further support the validity and robustness of our GWAS findings. However, differences are noted in the upper tail of the p-value distribution, where the maximum observed –log10(p) values vary: 82.3* 10^-8^ or the All-controls, 72.5 * 10^-8^ for the Matching, and 66.1* 10^-8^ for the Random.

**Figure 2.**
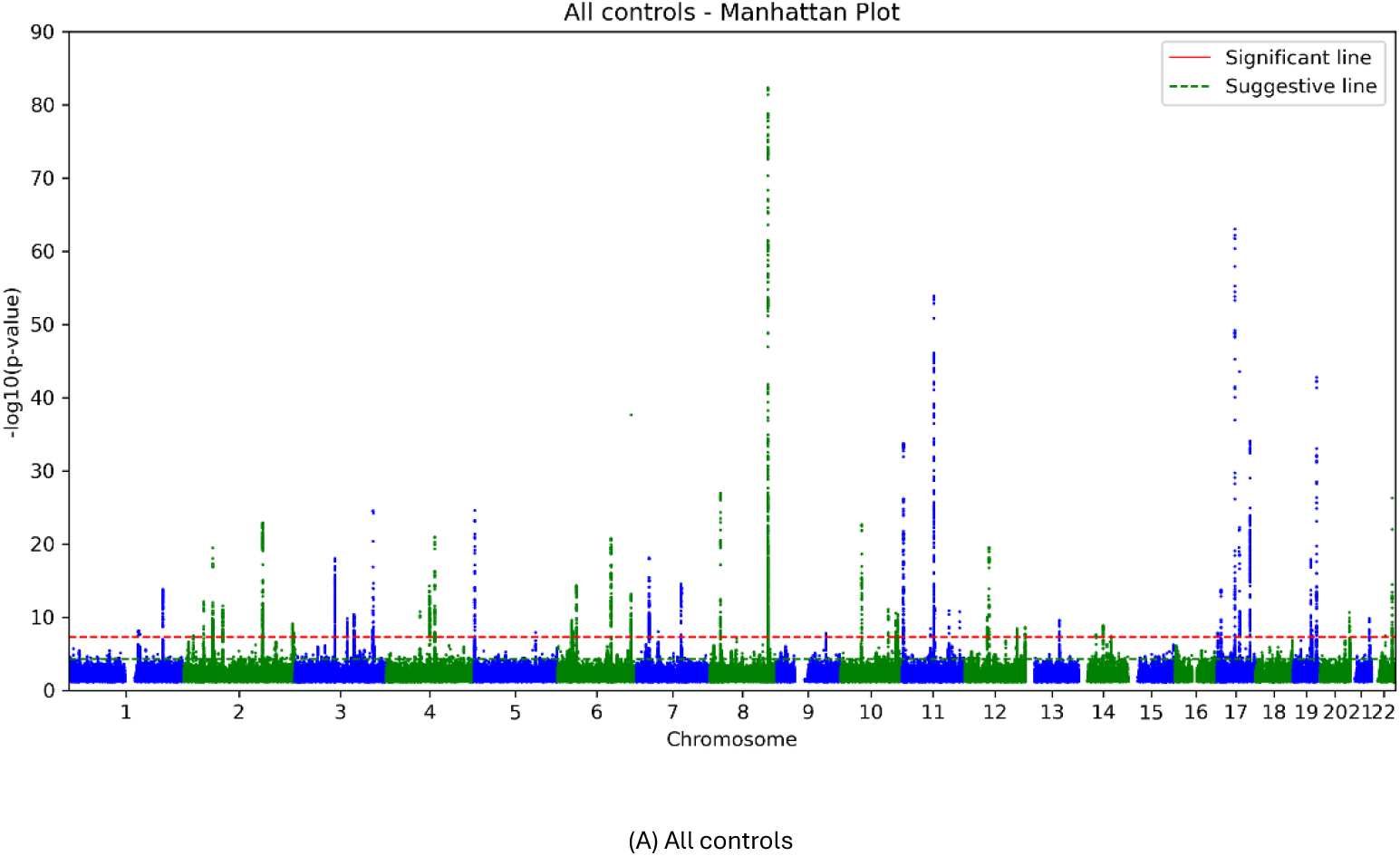
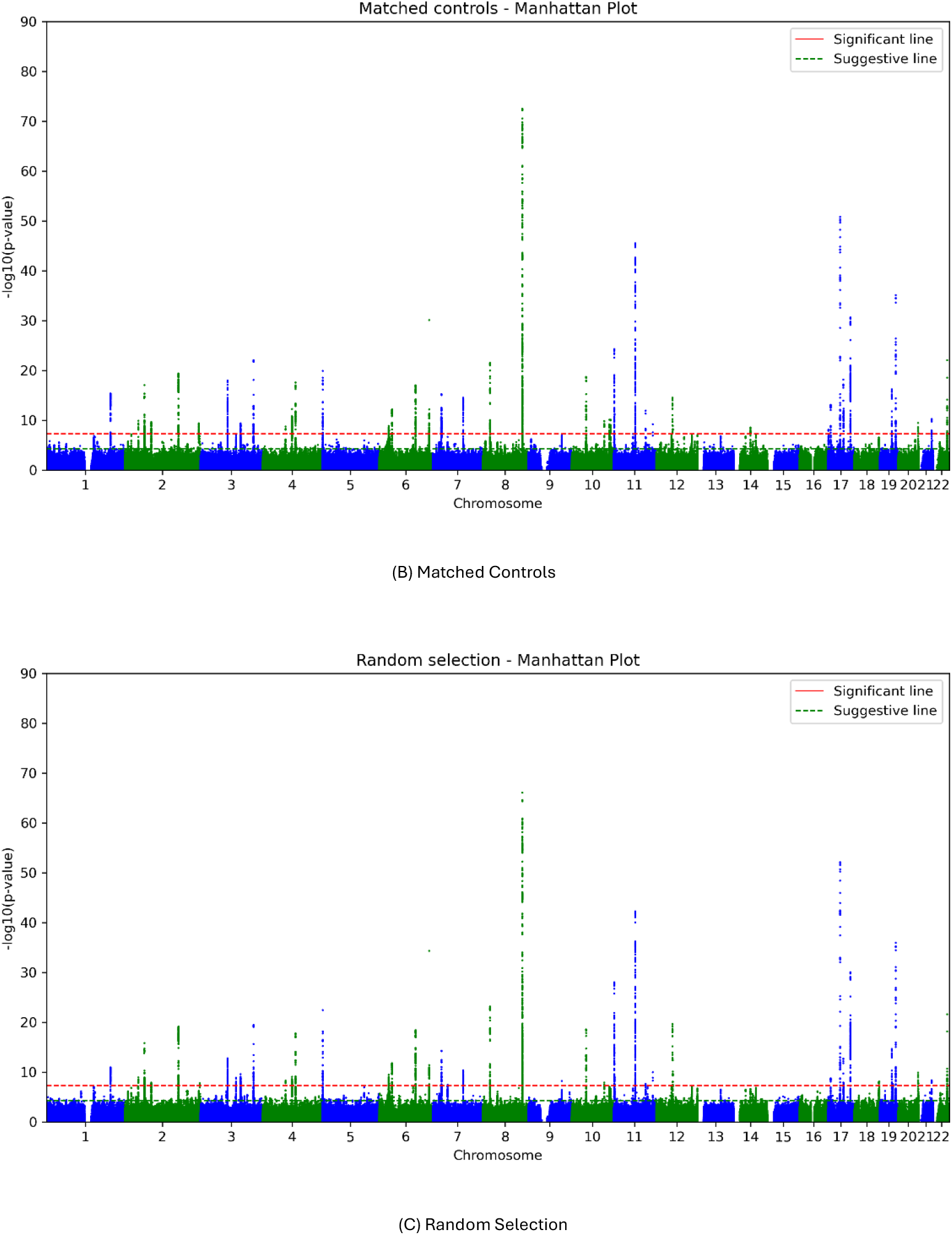
Manhattan plots of GWAS results under different control strategies. Overall signal patterns are consistent, while the top – log10(p) peaks differ at 82.3, 72.5, and 66.1.

**Figure 3.**
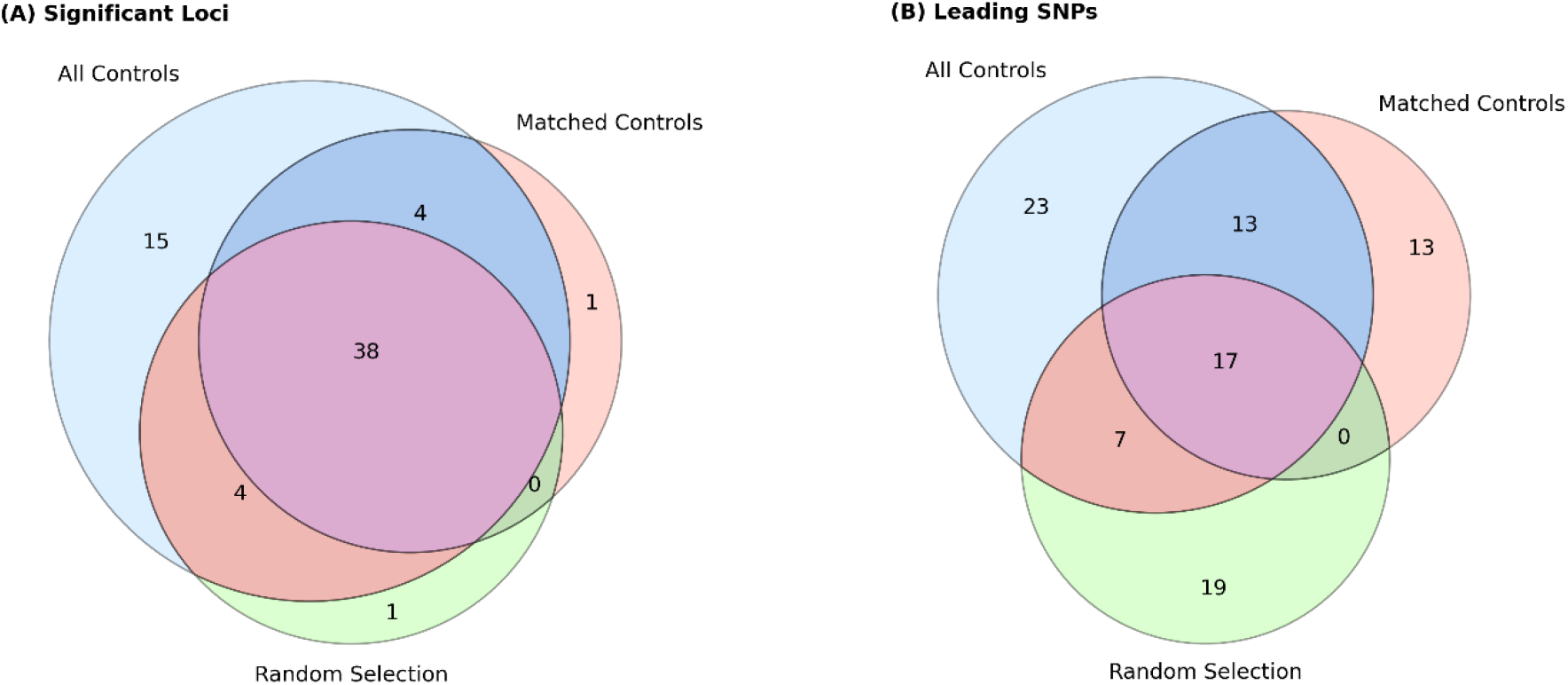
Venn diagram of significant loci and leading SNPs across GWAS from All, Match and Random cohorts.

### 3.3 GWAS Significant and suggestive SNPs

We compared the number and concordance of genome-wide significant (P < 5 × 10−8) and suggestive (P < 5 × 10−5) SNPs identified under three strategies: All, Match, and Random (**Table 2**). The total number of tested SNPs varied slightly across strategies due to control number differences, with All-controls analysing over 13.5 million SNPs, compared to ∼10.9 million in Match and Random.

**Table 2.** GWAS results of significant and suggestive SNPs statistics across three control strategies.

| Strategies | Total SNP Numbers | Max $-\log_{10}(P)$ | Percentages of SNPs |
| --- | --- | --- | --- |
| **All Controls** | **13,513,780** | **82.34** |  |
| *Significant | 4,572 | | $3.4 \times 10^{-4}$ |
| *Suggestive | 11,329 | | $8.4 \times 10^{-4}$ |
| **Match Controls** | **10,910,234** | **72.50** |  |
| Significant | 3,572 | | $3.3 \times 10^{-4}$ |
| Suggestive | 9,233 | | $8.5 \times 10^{-4}$ |
| **Random Selection** | **10,875,211** | **66.10** |  |
| Significant | 3,064 | | $2.8 \times 10^{-4}$ |
| Suggestive | 8,503 | | $7.8 \times 10^{-4}$ |

\*Significant threshold: $P < 5 \times 10^{-8}$ ; Suggestive threshold: $P < 5 \times 10^{-5}$

While All and Match yielded comparable capture rates for both significant and suggestive SNPs, Random exhibited a noticeably lower yield in both categories. Specifically, All identified 4,572 significant SNPs (3.4×10^-4^ ) % and 11,329 suggestive SNPs (8.4×10^-4^) %; Match identified 3,572 significant (3.3×10^-4^) % and 9,233 suggestive (8.5×10^-4^) % SNPs; and Random yielded 3,064 significant (2.8×10^-4^) % and 8,503 suggestive (7.8×10^-4^) % SNPs.

### 3.4 GWAS locus signal results

The **Table 3** displays genome-wide significant association signals (–log10P > genome-wide threshold) detected across three control selection strategies (All, Matched, Random). Shown are loci restricted to chromosomes 8, 10, 11, and 17, which had the most significant findings across the three analyses. Each row represents the lead SNP per locus with –log10P values and allelic information.

**Table 3.** GWAS significant loci with leading SNPs and P values on chromosomes 8, 10, 11, and 17.

| Chr | All leading SNP | Match leading SNP | Random leading SNP | All P value * | Match P value | Random P value |
| --- | --- | --- | --- | --- | --- | --- |
| 8 | 23665232<br>A/G |  | *23668030<br>T/C | 26.92 | 21.59 | 23.19 |
| 8 | 127516190<br>C/G | *127518371<br>G/A | *127064901<br>A/G | 82.34 | 72.5 | 66.10 |
| 10 | 46098665<br>C/CA |  | *46099199<br>C/A | 22.67 | 18.73 | 18.58 |
| 10 | 103908421<br>CTT/C |  |  | 11.09 | 9.86 | 7.91 |
| 10 | 121038900<br>G/A |  |  | 10.59 | 10.11 |  |
| 10 | 125008517<br>A/G |  |  | 10.37 | 10.13 |  |
| 11 | 2211394<br>C/T | *2212255<br>G/C | *2207388<br>C/T | 33.74 | 24.30 | 28.04 |
| 11 | 62140968<br>C/T |  |  | 8.43 | - | - |
| 11 | 69236512<br>T/A |  |  | 53.91 | 45.54 | 42.28 |
| 11 | 102530930<br>C/T |  |  | 10.93 | 11.96 | 7.69 |
| 11 | 125274530<br>G/A |  |  | 10.77 | 9.20 | 9.99 |
| 17 | 715725<br>C/T | *714799<br>A/G |  | 7.85 | 8.11 | - |
| 17 | 7913036<br>G/T |  |  | 13.74 | 13.09 | 8.78 |
| 17 | 37743574<br>G/A |  | *37739961<br>T/A | 63.04 | 50.84 | 52.13 |
| 17 | 48728343<br>T/C | *48612699<br>A/AGAGAGC | *48654889<br>T/C | 43.55 | 18.20 | 12.75 |
| 17 | 71108797<br>G/A |  | *71120794<br>G/A | 34.09 | 30.62 | 30.05 |

\*Position: The difference leading SNPs located the same loci with all-controls; \* P value: $-\log(10)$ transformed p-value here

The divergence of both Match and Random strategies from the All-controls analysis was notable, with the largest discrepancy observed for random. Among the 60 loci identified using All-controls, approximately 30% leading SNPs were not replicated in the Match group and more than 50% leading SNPs were absent from the Random group. Furthermore, many loci showed attenuated test statistics or inconsistent effect directions in match and especially in random. For instance, lead SNPs at 8q24.21 (chr8:127516190; P = 8.2×10−8^3^) and 11q13.3 (chr11: 69236512; P = 5.4×10−^54^) remained significant in both All and Match, but the corresponding signals weakened or dropped below significance in Random, suggesting reduced sensitivity under random selection. Notably, despite Match and Random detecting a similar number of significant loci, Random often identified different leading SNPs compared to All-controls, indicating frequent lead SNP swapping under random selection.

A total of 60, 43, and 43 genome-wide significant loci were identified in GWAS using All, matched, and random, respectively. In **Figure 3**, shows 38 loci were significant across all three analyses. The overlap between All and Match, as well as between All and Random, was 42 loci in each case, indicating that both matched and random control designs achieved comparable sensitivity in detecting associated regions. However, analysis of leading SNPs revealed greater consistency between All and Match (30 shared SNPs), compared to All and Random (24 shared SNPs), suggesting that the matched design provided more precise localization of association signals. The smaller overlap between Match and Random compared to other pairings (17 shared SNPs and 38 loci, all of which were also shared with All) highlights divergent signal prioritization, implying that random controls may often select alternate tag SNPs within the same loci.

Among the unique leading SNPs to the match GWAS (n = 13), 11 (84.62%) were in high linkage disequilibrium (LD; r^2^ > 0.8) with SNPs identified in all GWAS. In contrast, only 11 out of 19 (57.89%) unique leading SNPs from the random GWAS showed such high LD with the all-controls SNPs, indicating improved signal consistency under the matched design. The detailed information of loci and leading SNPs provided in the supplementary table.

### 3.5 GWAS results show diff strategies OR similar with Conti’s results

The scatter plot of **Figure 4** illustrates the correlation between β estimates from Conti et al., (2021)’s study (x-axis) and the GWAS results under the “Matched controls” strategy (y-axis). Each point represents a genetic variant (MAF >1%). A linear regression line is overlaid, indicating strong concordance in effect size estimates between the two analysis. Similar linear relationships were observed when using All controls and Random selection strategies. Therefore, only one representative scatter plot (Matched controls vs. Conti) is shown. The adjusted R^2^ of 0.82 for the three linear regression models suggests that approximately 82% of the variance in GWAS effect sizes can be explained by Conti’s results, reflecting high consistency and supporting the notion that around 82% of observed GWAS signals in the real world are reproducible across different control strategies.

**Figure 4.**
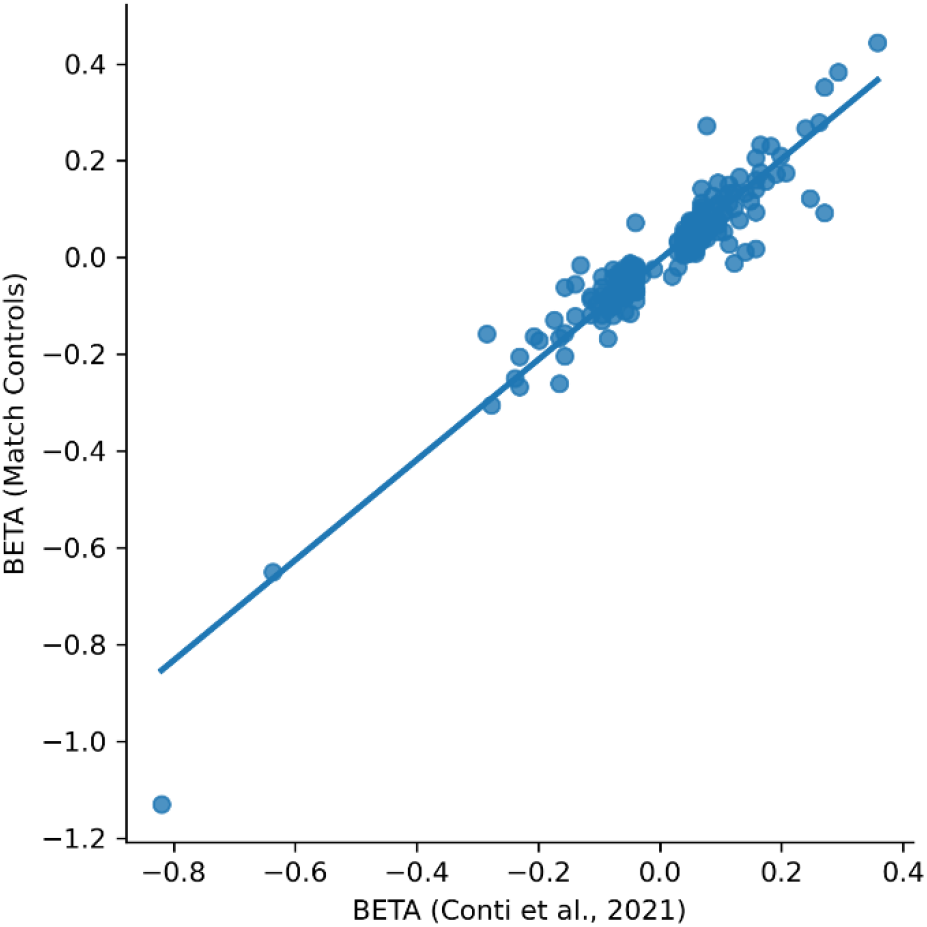
Concordance of 269 SNPs effect size estimates between Conti’s study and GWAS with Matched Controls.

## 4 Discussion

### 4.1 Principal Findings

This study evaluated how control selection strategies influence GWAS outcomes. We compared three distinct approaches: All, Matched, and Random. While all strategies revealed broadly similar global association patterns, reflected in the overall alignment of Manhattan plots and identification of key prostate cancer loci, the depth and consistency of association signals varied notably across designs.

A similar percentage of genome-wide significant SNPs were identified between all controls 3.4×10^-4^ (0.034%) and matched controls 3.3×10^-4^ (0.033%), while using random controls identified a significantly lower percentage of SNPs 2.8×10^-4^ (0.028 %). While both Matching and Random selection resulted in a ∼30% reduction in genome-wide significant loci compared with All-controls, Matching offered greater consistency with All-controls in terms of leading SNP identification and direction of β estimation.

### 4.2 Interpretation

Our findings demonstrate empirically the trade-off between computational intensity and statistical power. The increased computational requirements of modern GWAS studies (especially when WGS data is used) incurs an increased computational cost. In our study, the GWAS with All-controls was performed with a total sample size of 200,000 male UK Biobank participants, while Random and Matching was performed with 60,000.

Optimal case-control ratios have long been studied in the context of genetic association studies. While increasing the number of controls can enhance statistical power, especially when case numbers are limited, the benefit of adding more controls diminishes beyond a certain point. Empirical and theoretical evaluations suggest that power gains plateau around a 1:4 to 1:5 case-control ratio, after which additional controls yield minimal improvement relative to cost and computational burden (Lakens, 2022; Ritchie et al., 2015).

In large-scale studies, controlling for unbalanced case-control ratios and accounting for relatedness among samples becomes computationally intensive. Moreover, the risk of population stratification and residual confounding tends to increase with larger sample sizes, as subtle structure in the data can lead to spurious associations (Turner et al., 2011; Uffelmann et al., 2021). This underscores the importance of careful control selection. Matching cases and controls on important covariates such as age, ancestry, and study centre can effectively reduce confounding due to unaccounted population structure and environmentally driven allele frequency differences (Yang et al., 2011; Zhou et al., 2020).

The utility of case-control matching becomes even more evident in studies of rare diseases. For traits with low prevalence, the ratio of cases to available controls can be extremely imbalanced, often 1:1000 or higher. While using all available controls may seem statistically appealing, such extreme imbalance increases computational burden, especially in models testing rare variants or modest effect sizes (Lee et al., 2014).

### 4.3 Strengths and Weaknesses

The matching on case-control selection is a plug-in approach to GWAS, which is easy to implement and combine with any pipeline or code module. Demographic comparisons show that matched controls more closely resemble cases in age and genetic background, improving covariate balance and reducing confounding. This is especially beneficial in large-scale whole genome (WGS) or exome sequencing (WES) studies, where computational and financial costs increase with sample size.

Although both the Match and Random groups are subsets of the overall All-controls group, they represent distinct sampling strategies. Consequently, the greatest divergence in association results is observed between the Match and Random designs. This divergence suggests that case-control matching more effectively controls for confounding, potentially yielding more reliable SNP-level associations compared to random selection. However, this improvement in internal validity comes at a cost. matching reduces the effective sample size by discarding unmatched controls, but it does not eliminate bias, which may lead to a loss of statistical power compared to using the full set of available controls.

This study has several limitations. First, prostate cancer cases were defined using ICD-10 codes in the UK Biobank, which lack histopathological detail and staging information. This may lead to phenotype misclassification and reduce the precision of genetic association estimates. Second, analyses were limited to individuals of European ancestry, which improves internal validity but restricts the generalisability of findings to other populations. Third, the study focused exclusively on prostate cancer as a representative phenotype. Although this enabled detailed comparison of control selection strategies, future studies should include a wider range of traits, particularly rare diseases and phenotypes with different genetic architectures.

### 4.4 Conclusion

From a practical perspective, we recommend case-control matching GWAS as an efficient strategy for rapid association testing across multiple phenotypes. This approach is particularly useful when the goal is to verify known loci or replicate associations in costly sequencing contexts, rather than to discover novel variants. Looking forward, we plan to extend these evaluations to WGS and WES data and to perform external validation in deeply phenotype cohorts such as the All of Us Research Program. This will allow us to assess generalisability across diverse ancestries and health systems.

## Data Availability

Availability and Implementation: R code for implementing matching and random control selection is provided and available on GitHub.

https://github.com/Jingzhan-Lu/GWAS-Control-Selection

## Acknowledgements

This study was supported by the National Institute for Health and Care Research Exeter Biomedical Research Centre (NIHR Exeter BRC) and and the University of Exeter Medical School. This research has been conducted using data from UK Biobank (http://www.ukbiobank.ac.uk/), a major biomedical database, and was made possible through access to the data and findings generated by the UK Biobank project number 103356.

## Notes

### Competing Interest Statement

The authors have declared no competing interest.

