## Supplementary Material for "Impact of control selection strategies on GWAS results: a study of prostate cancer in the UK Biobank"

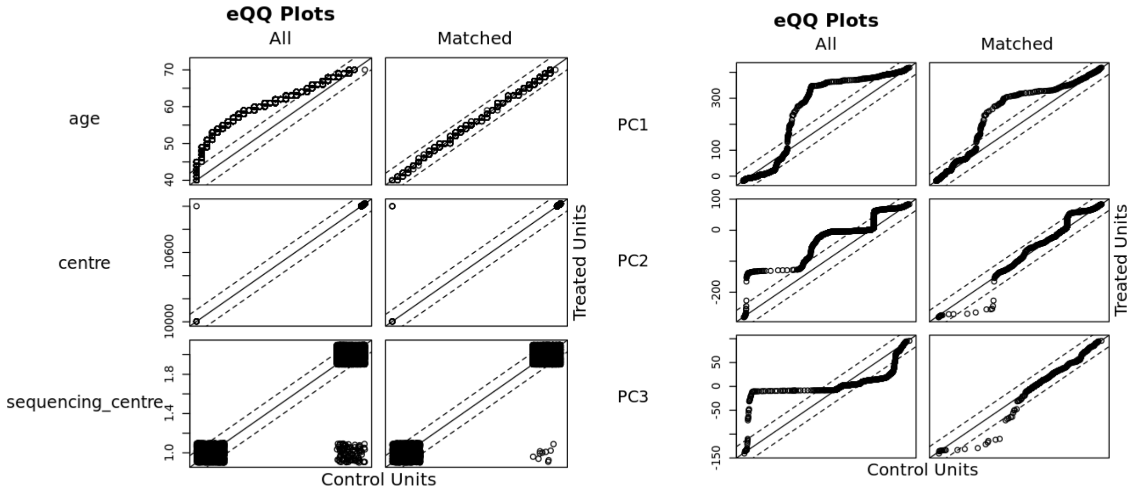


**Supplementary Figure 1.** Covariates balance assessment using eQQ Plots Before and After Matching

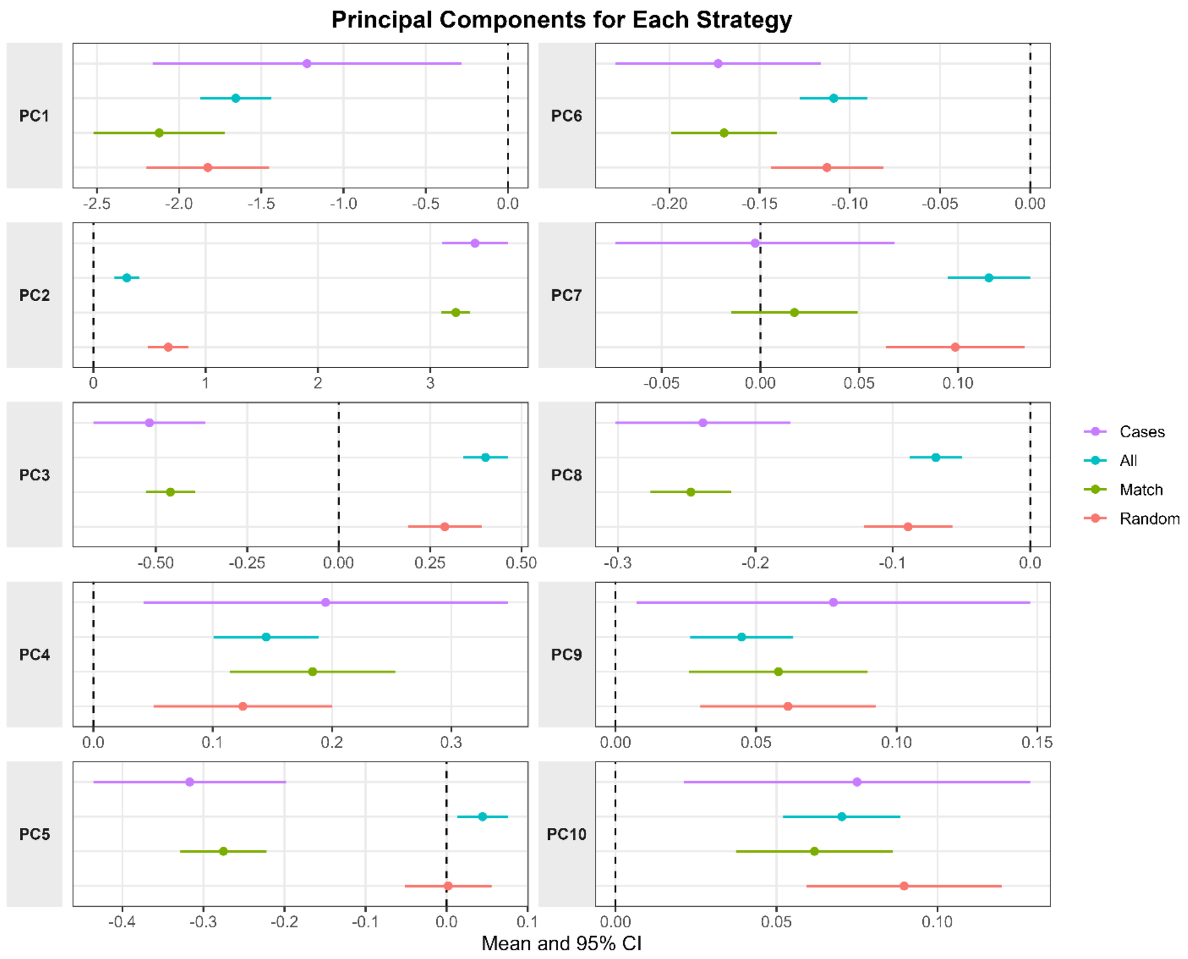


**Supplementary Figure 2.** Comparison of the Top 10 Principal Genetic Components for each strategy

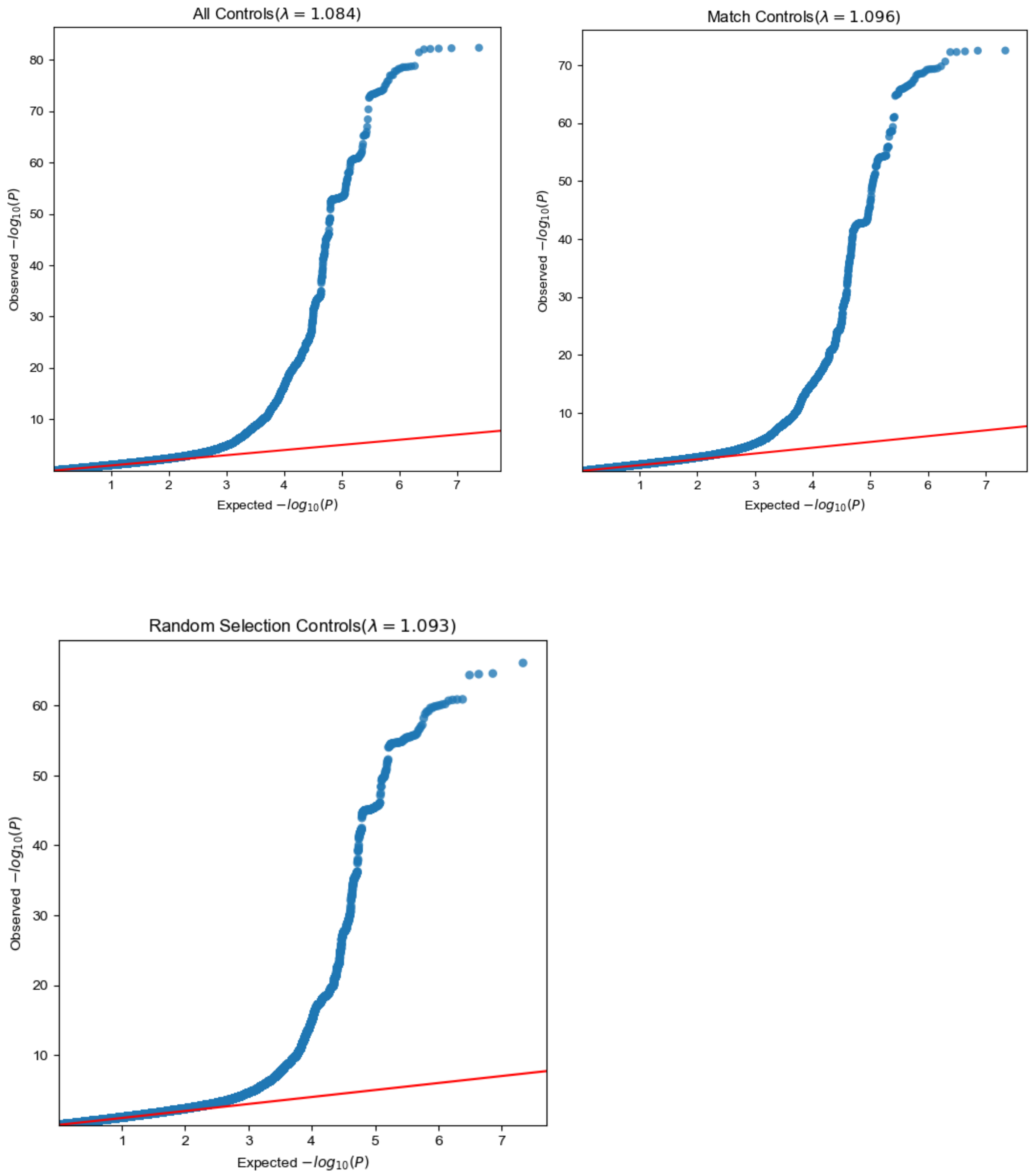


**Supplementary Figure 3.** QQ plots and genomic inflation factors (λ) for different control selection strategies.

**Supplementary Table 1.** GWAS Sign Test across All, match and random strategies

| Sign Test | ++ (positive) | -- (negative) | +-/-+ (different direction) |
|---|---|---|---|
| S1 vs S2 (All vs Match) | 55.44% | 44.50% | 0.06% |
| S1 vs S3 (All vs Random) | 57.64% | 42.33% | 0.03% |
| S2 vs S3 (Match vs Random) | 56.46% | 43.50% | 0.04% |
